# Spatial characterization and stratification of colorectal adenomas by Deep Visual Proteomics

**DOI:** 10.1101/2023.11.25.23298987

**Authors:** Sonja Kabatnik, Frederik Post, Lylia Drici, Annette Snejbjerg Bartels, Maximilian T Strauss, Xiang Zheng, Gunvor Iben Madsen, Andreas Mund, Florian A Rosenberger, José MA Moreira, Matthias Mann

**Author notes:** Correspondence: **Matthias Mann**, PhD, Max-Planck Institute of Biochemistry, am Klopferspitz 18, 82152 Martinsried, Germany.; or **José Manuel Afonso Moreira**, PhD, Department of Drug Design and Pharmacology, Faculty of Health and Medical Sciences, University of Copenhagen, Universitetsparken 2, DK-2100 Copenhagen, Denmark.

## Abstract

Colorectal adenomas (CRA) are precursor lesions of the colon that may progress to adenocarcinomas, with patients currently categorized into risk groups by morphological features. We aimed to establish a molecular feature-based risk allocation framework towards improved patient stratification. Deep Visual Proteomics (DVP) is a novel approach that combines image-based artificial intelligence with automated microdissection and ultra-high sensitive mass spectrometry. Here we used DVP on formalin-fixed, paraffin-embedded (FFPE) CRA tissues from nine patients, immunohistologically stained for Caudal-type homeobox 2 (CDX2), a protein implicated in colorectal cancer, enabling the characterization of cellular heterogeneity within distinct tissue regions and across patients. DVP seamlessly integrated with current pathology workflows and equipment, identifying DMBT1, MARCKS and CD99 as correlated with disease recurrence history, making them potential markers of risk stratification. DVP uncovered a metabolic switch towards anaerobic glycolysis in areas of high dysplasia, which was specific for cells with high CDX2 expression. Our findings underscore the potential of spatial proteomics to refine early-stage detection and contribute to personalized patient management strategies and provided novel insights into metabolic reprogramming.

## Introduction

Colorectal cancer (CRC) is one of the most prevalent cancers, with a poor prognosis when detected at later stages. Worldwide, it is the third most diagnosed cancer and the second-leading cause of cancer deaths. Two thirds of CRC cases arise sporadically by a combination of multiple environmental risk factors, such as unhealthy diet, physical inactivity, obesity, smoking and excessive alcohol consumption (Morgan et al., 2022; Siegel et al., 2020).

CRC screening programs, together with removal of precancerous lesions – colonic adenomatous polyps (adenomas) – have led to decreasing incidence rates in older adults in high-income countries, whereas incidence is rising in developing countries, at least for adults under 50 years (Morgan et al., 2022). Regular screenings strongly decrease CRC-associated mortality because detected adenomas are removed, followed by scheduled follow-up screening based on a set of criteria, including the number of adenomas removed, their size and histopathologic presentation (Hassan et al., 2020). The presence of high-grade (HG) dysplasia categorizes a precursor lesion as high-risk, requiring patients to undergo regular endoscopic examinations. Given that only 5-10% of HG dysplasia adenomas will ever develop into carcinomas, this leads to overtreatment and frequent surgical removals of nonmalignant colorectal polyps, a burden for both the individual and the healthcare system (Le Roy et al., 2015; Neugut et al., n.d.; Peery et al., 2018). It is therefore important to develop a more sophisticated and robust workflow for CRA classification that is supported by quantitative molecular data rather than only IHC and cell morphology.

Tissue samples collected for diagnostic purposes are routinely preserved by formalin-fixation and paraffin-embedding (FFPE) which can be archived in hospitals for decades. These fixed and embedded tissue samples exhibit remarkable stability, allowing for their extended use in clinical histopathological assessments, and offering the potential for continued research utilization many years after their initial preparation (Grillo et al., 2017). However, different macromolecules are affected differently by storage time and conditions (Greytak et al., 2015; Yakovleva et al., 2017).

In recent years, MS-based proteomic technologies have improved markedly in sensitivity and robustness, now routinely allowing large-scale analysis of FFPE tissues in a clinical context (Coscia et al., 2020). This was enabled by advances in sample preparation protocols allowing highly efficient peptide recovery, robust and streamlined liquid chromatography (LC) set ups and increasingly powerful mass spectrometry (MS) instruments (Bache et al., 2018; Brunner et al., 2022; Coscia et al., 2020). Furthermore, data independent acquisition (DIA) modes such as diaPASEF (parallel accumulation-serial fragmentation) have enabled high ion utilization on quadrupole-TOF (time-of-flight) mass analyzers, yielding high data completeness even for low-input samples (Brunner et al., 2022; Meier et al., 2020). Very recently, Deep Visual Proteomics (DVP) has made it possible to analyze collections of the same single-cell types or states (Mund et al., 2022). DVP utilizes high-resolution image information, combines it with automated single-cell laser microdissection and ultra-high sensitivity mass spectrometry (MS) and is readily applied to FFPE material.

In this work, we set out to develop a robust and streamlined DVP pipeline that could enable a spatially resolved, in-depth molecular characterization and prognostic stratification of CRAs. We selected Caudal-type homeobox transcription factor 2 (CDX2) as the guiding feature for our DVP analysis of CRAs. CDX2 is a key regulator of intestinal differentiation and homeostasis (Badia-Ramentol et al., 2023). In CRC, CDX2 functions either as a tumor-suppressor gene (Aoki et al., 2003; Balbinot et al., 2018; Bonhomme et al., 2003) or as an oncogene (Salari et al., 2012), depending on context. In addition, CDX2 regulates immune cell infiltration in the intestine, modulating local immune responses (Dalerba et al., 2016a; San Roman et al., 2015). Loss or decreased expression of CDX2 is a common event in CRC, associated with molecular features such as CpG island methylator phenotype, and microsatellite instability(Baba et al., 2009) and may be predictive of a more aggressive disease course (Baba et al., 2009; Dalerba et al., 2016b). The need to clarify the mechanistic role of decreased CDX2 expression in the pathogenesis of colon cancers, together with the fact that mutations in the CDX2 gene are extremely rare in CRC (Hinoi et al., 2003). make this protein well suited to the discovery of tumor stratification markers using DVP.

We collected HG dysplasia adenoma tissue samples from nine individuals based on their clinical history and divided them into three groups: development of either metachronous CRC (C), metachronous HG adenoma (HDA) within five years, or no new lesions at least up to 10 years of colorectal surveillance, categorized as the group of non-metachronous neoplasms (NMN), and describe here the results of our CDX2-guided spatially resolved, in-depth proteomic analysis of these samples.

In conclusion, our optimized DVP workflow readily integrated into well-established clinical protocols, seamlessly aligning with existing routine pathology practices. Our study furnished comprehensive spatial proteome data at the single-cell type level, addressing the inherent heterogeneity that is intrinsic both within individual tumors and among different patients (Bedard et al., 2013; Marusyk et al., 2020). This unveiled novel perspectives on region-specific protein landscapes, shedding light on biologically significant factors and spatially localized processes that could play pivotal roles in CRA classification and clinical decision-making.

## Results

### Study design and the Deep Visual Proteomics workflow

To investigate molecular protein signatures associated with disease progression in CRA, we employed a retrospective study design that comprised a cohort of male individuals who had undergone polypectomy procedures for CRA removal (Bech et al., 2023). We selected a sub-cohort of nine patients that all had high-grade (HG) dysplasia adenomas as verified by a pathologist using morphological criteria and immunohistochemical (IHC) analysis, but had heterogeneous clinical outcomes. Of the nine patients, three were diagnosed with metachronous colorectal cancer (C) within a year, three developed new lesions exhibiting HG dysplasia characteristics once again (HDA), while the remaining three patients remained free of new neoplastic lesions throughout the entire 10-year surveillance duration (NMN) (Table EV1).

Note that the initial diagnosis of HG dysplasia in adenomas uniformly mandated the scheduling of costly yearly follow-up surveillance colonoscopies, a measure that, while essential for patient care, may pose practical challenges such as poor patient compliance as well as burdening the health care system (Figure 1A). This highlights the desirability of complementing existing pathology practices with molecular data.

**Figure 1.**
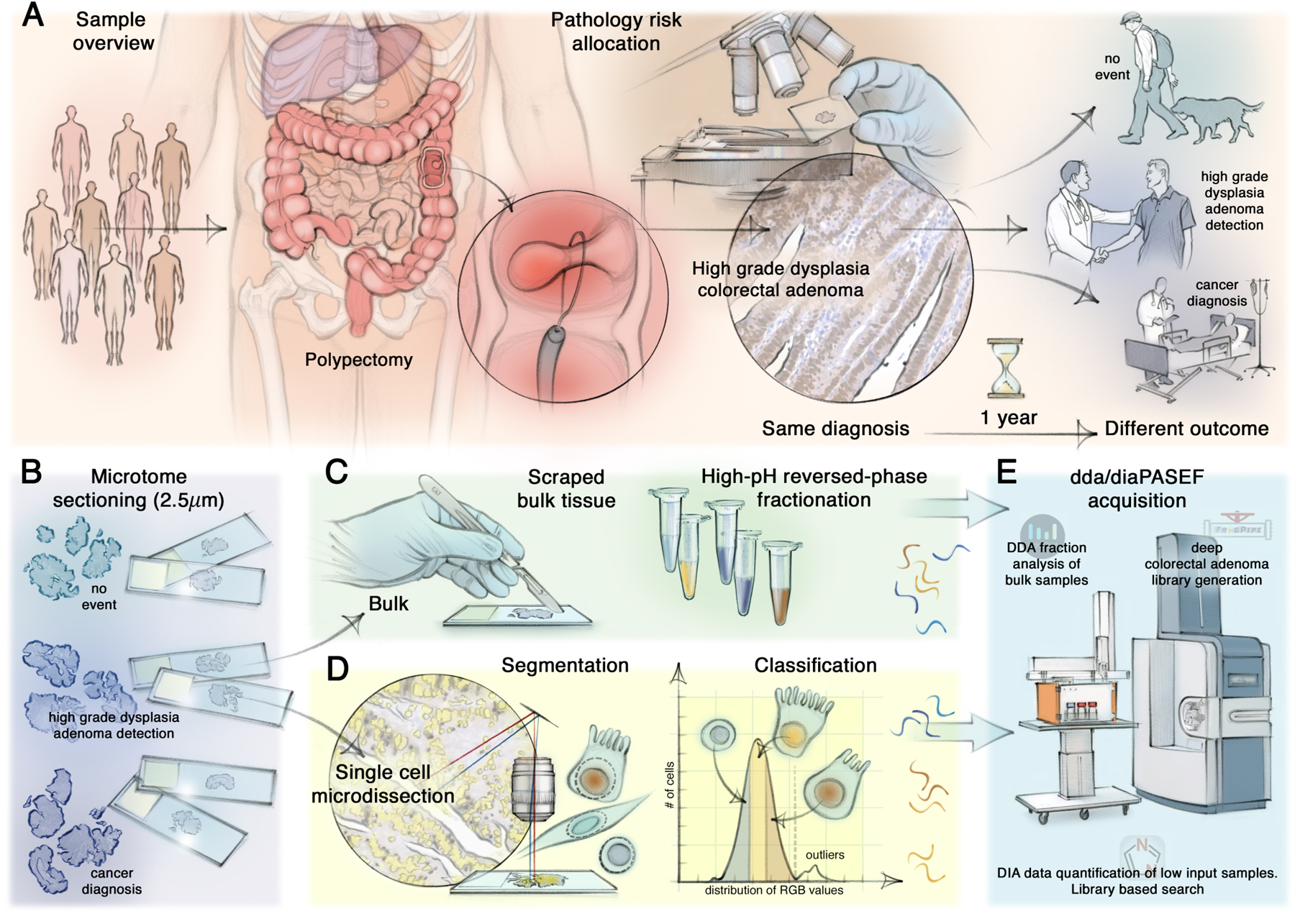
Study design and our multi-layered mass spectrometry-based proteomics approach. A. Schematic representation of the colorectal adenoma (CRA) cohort and the study design. Nine resected CRAs displayed high-grade (HG) dysplasia which led to the same pathological assessment and diagnosis but showed three different clinical outcomes. B-E. Multi-layered and streamlined mass spectrometry (MS)-based proteomics approach applied to FFPE CRA tissues. B. FFPE blocks of each polyp were cut and mounted onto PEN membrane slides. C. For bulk proteomics analysis, each tissue was scraped, lysed, digested, and extensively fractionated. D. Deep Visual Proteomics (DVP) workflow for the analysis of region and cell class-specific protein changes, including machine learning (ML)-based segmentation, RGB and morphology-dependent classification, followed by automated laser microdissection. E. Bulk proteomics and low-input DVP samples were measured on the same EvoSep-timsTOF platform, either in data-dependent (ddaPASEF) or data-independent (diaPASEF) acquisition mode, followed by spectral identification and quantification with AlphaPept, MSFragger or DIA-NN.

To this end, we applied a high-throughput MS workflow which we streamlined to measure our unique cohort of nine non-malignant CRA FFPE samples, all of which were more than eleven years old. Building on standardized IHC pathology protocols and markers, we mounted each sample onto PEN membrane microscopy glass slides in duplicates and either performed bulk proteomics in DDA and DIA mode, or cell-type specific DVP (Figure 1B). For bulk analysis, we left the mounted FFPE tissues unstained and scraped them from the slides for lysis, protein digestion and subsequent extensive reverse-phase, high pH fractionation of the resulting peptides (Figure 1C, Materials and Methods).

To gain insight into spatially resolved protein signatures of specific cell classes, we stained each CRA tissue for CDX2 and counterstained with hematoxylin. We combined simple IHC staining and widefield image acquisition, which we coupled to a machine learning-based nucleus segmentation model. We then classified cells based CDX2 staining. After outlier elimination, and segmented contours were categorized into three classes based on normalized RGB intensities, ratios, and form features (CDX2++, CDX2+ and CDX2-, Figure 1D). Class CDX2++ and CDX2+ were epithelial colon cells, characterized by high and medium marker expression, respectively, while class CDX2-were stromal.

We excised 1,000 shapes per class in triplicates (corresponding to about 100 complete cells in these thin sections) by automated laser microdissection, and collected those into a 384-well plate (Figure 1D). Using the EvoSep One chromatography system and the 30 Samples Per Day method (“30 SPD”), we were able to robustly measure hundreds of FFPE-derived bulk and low-input samples on our timsTOF mass spectrometer (Figure 1E)

To obtain an in-depth proteome overview for each patient, we acquired fractions in DDA mode and quantified them using AlphaPept (Strauss et al., 2021), our open-source Python-based MS search engine. Additionally, we employed these files to construct a very extensive CRA library by the FragPipe computational proteomics platform with integrated MSFragger (Kong et al., 2017a; Yu et al., 2020). In contrast, all low-input DVP samples were acquired in DIA mode and searched using the DIA-NN against our comprehensive, project-specific CRA library (Demichev et al., 2022) (Figure 1E, Material and Methods).

### A very deep proteomics resource of non-malignant colorectal adenomas

To obtain maximal proteomic depth from our archival, room temperature stored FFPE CRA tissues, we applied our DVP sample preparation protocol, including extensive reverse-phase, high pH fractionation, resulting in 48 fractions per patient and 432 samples in total. We measured these in just 14 days on our liquid chromatography system, coupled to our ultra-high sensitivity mass spectrometry instrument (Material and Methods). Median protein depth across samples was nearly 4,000 unique proteins, summing up to deep CRA library of 12,380 proteins from 178,274 unique, identified peptides, all at an FDR of 1% (Figure 2A). Given that there are about 20,000 protein coding genes, this constitutes excellent proteomic coverage, which was also supported by identification of 79% of TARGET (tumor alterations relevant of genomics-driven therapy) database-annotated genes known to be associated with disease progression, CRC, and resistance (Figure 2B) (Armaghany et al., 2012; Van Allen et al., 2014). We then selected the maximum intensity value for each protein across the fractions of each patient for differential analysis, which yielded a median protein number of more than 10,000 for each of the patients (Figure 2C). We expected the largest biological variation between those that had developed cancer and those that with no neoplasms after 10 years ( C and NMN), and performed a two-tailed student’s t-test with multiple-hypothesis testing correction (premutation-based FDR <0.05, s_0_ = 0.1) between them. However, this analysis resulted in only minor differential expression, possibly because of the small number of patients and the relatively high Coefficient of Variation (CV), caused by the extensive fractionation procedure (Figure EV1A).

**Figure 2.**
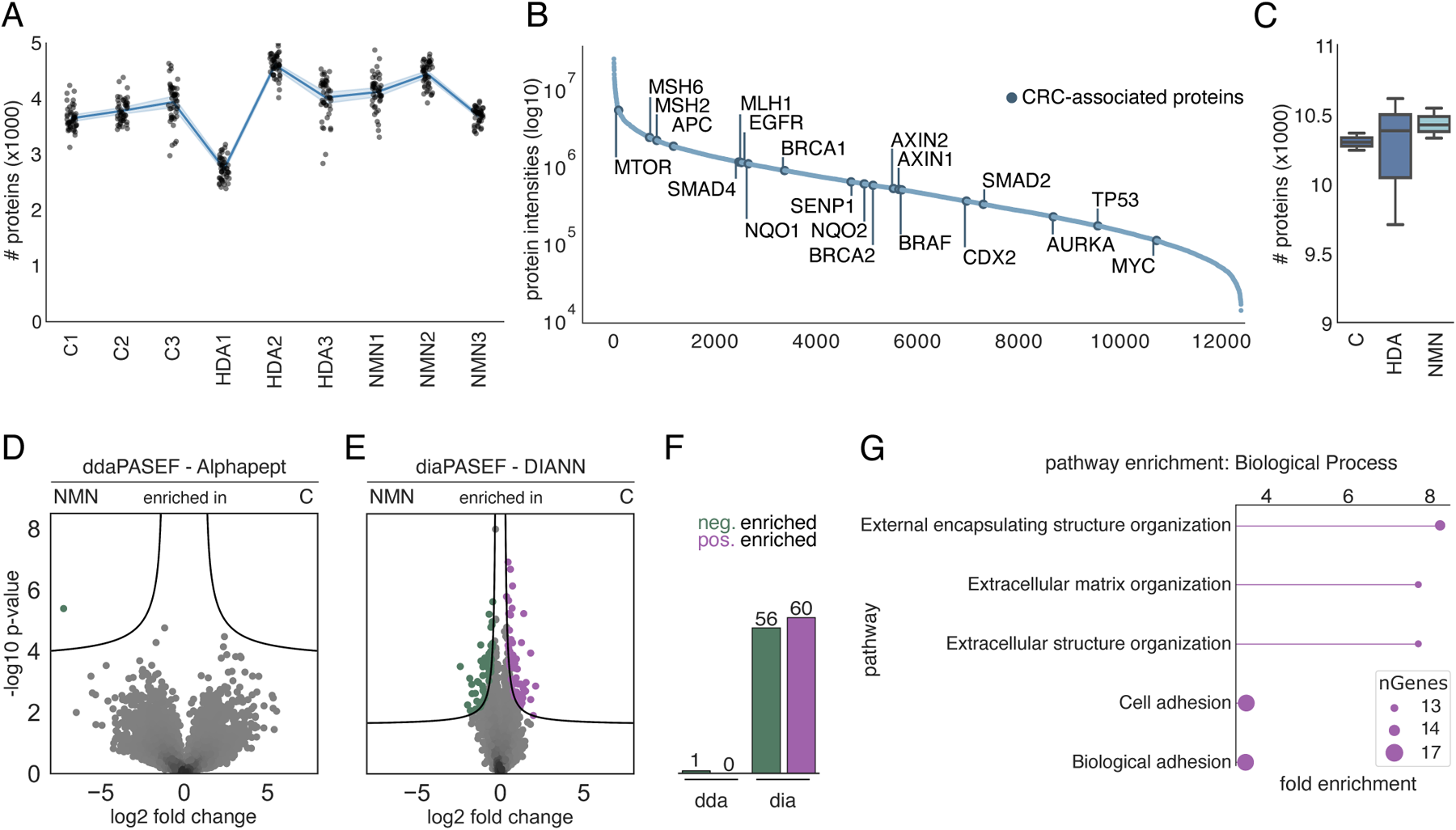
Bulk FFPE tissue proteomics of non-malignant colorectal adenomas. A. Number of proteins in each DDA-acquired fraction per CRA sample. B. Normalized total intensity of all identified proteins in our deep CRA library created from 432 fraction samples. Highlighted in dark blue: colorectal cancer (CRC)-associated proteins, part of the TARGET (tumor alterations relevant of genomics-driven therapy) database. C. Number of proteins per patient within a group. D, E. (D) Pairwise proteomic comparison between C and NMN patient adenoma samples, acquired in DDA, or (E) DIA mode. DDA data originated from fractionated samples, DIA was measured as single run (50 ng). Significantly enriched proteins are colored and displayed above the black lines indicating statistical significance (two-sided t-test, premutation-based FDR <0.05, s_0_ = 0.1). F. Number of significantly down- and upregulated proteins in the volcano plot analyses. G. GO Biological Process enrichment (FDR <0.05) of significantly upregulated protein hits.

Next, we measured unfractionated lysate for each patient directly in the DIA mode using the deep DDA library for matching. Supporting our conjecture, this resulted in many more statistically significantly regulated proteins (60 upregulated in the cancer group and 56 upregulated in the NMN group) (Figure 2E and F). Gene Ontology (GO) term enrichment on the upregulated proteins in DIA data revealed significant effects on processes related to extracellular structure reorganization and cell-cell adhesion (Figure 2G). Moreover, Gene Set Enrichment Analysis (GSEA) identified the involvement of RAF1, oncogenic MAPK signaling and mitochondrial translation pathways (Figure EV1B). This finding reflects augmented proliferation and cell migration within CRAs of group C, alongside a metabolic shift favoring anaerobic glycolysis (Ehrenreiter et al., 2005; Koc et al., 2022). Although this proteomics analysis constitutes positive control as it largely recapitulates known features of CRC, it clearly suffers from the limitation of bulk tissue analysis, in terms of spatial resolution and in assigning these effects to specific cell types, pointing to the desirability of DVP analysis. As a preliminary step, we next analyzed down to 5 ng of FFPE bulk tissue, an amount that would readily be available in DVP. Encouragingly, we found similar protein signatures in these experiments (Figure EV1C and D).

### Characterization of the colorectal adenoma landscape by DVP

Before applying our spatial DVP workflow across the different cell types in the CRA cohort, we first explored the protein landscape of the disease in one individual. We chose a particularly heterogeneous adenoma that was surgically removed from the colon of a male individual (designated C3 in Table EV1). In the following year, this patient had been diagnosed with CRC upon surveillance follow-up colonoscopy. For this case study, we stained the tissue by IHC against CDX2 as a marker for high grade dysplasia that is associated with colorectal tumorigenesis. Based on expression of this marker and tissue morphology in a widefield image, we chose three tissue areas which are characterized by: focal high dysplasia (region 1, purple); low dysplasia (region 2, yellow); as well as a region with normal glandular architecture and strong lymphocyte infiltration (region 3, green) (Figure 3A and B, Figure EV2A). Quantifying the distribution of classified cells across the whole tissue and annotated areas, we found that 52% of all CDX2++, and 30% of CDX2+ cells were located in the high dysplasia region. The proportion of CDX2-stroma cells, however, was similar in all regions (12-16%, Figure 3C).

**Figure 3.**
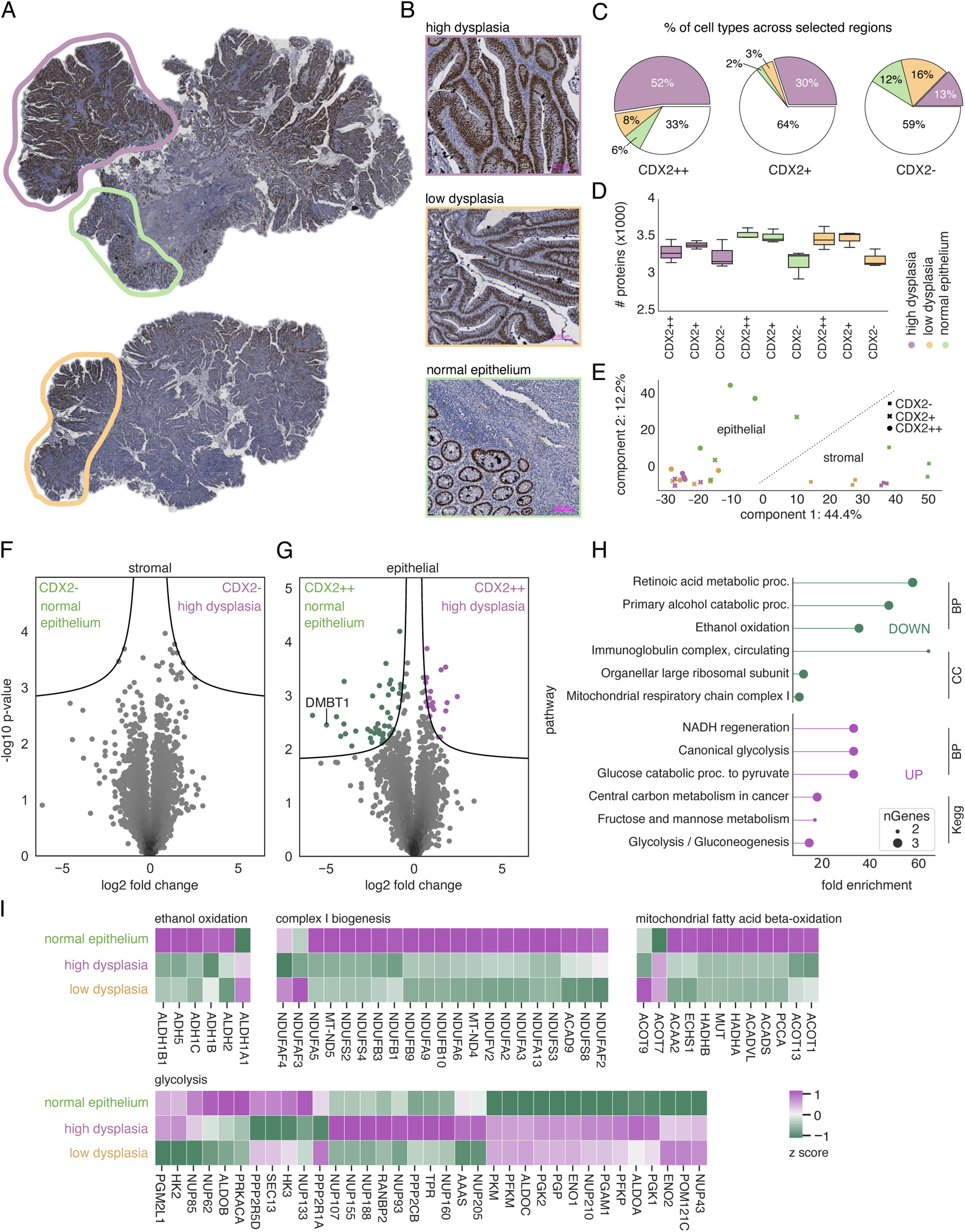
DVP characterizes region-specific metabolic changes within strongly heterogenous CRA sample. A. IHC staining of patient tissue C3 with three annotated tumor areas. B. Representative images of selected regions based on the degree of dysplasia, density of CDX2++ epithelial cells and lymphocyte infiltration (also see Supplementary Figure 3D). The color code signifies: 1, high dysplasia with a high density of CDX2++ cells (purple); 2, low dysplasia and medium density of CDX2++ cells (yellow); 3, normal glandular architecture and lymphocyte infiltration (green). Scale bar, 100 µm. C. Distribution of CDX2++, CDX2+ and CDX2-cells across annotated regions and the remaining whole tissue area (white). Note that percentages are rounded and may not add up to 100%. D. Unique protein numbers identified in CDX2++, CDX2+ and CDX2-cells. 1000 contours collected in triplicates. E. Principal component analysis (PCA) of collected CDX2++ and CDX2-cells across respective regions. F, G. Pairwise proteomic comparison of CDX2- and CDX2++ between the highly dysplastic area and the region with normal epithelium. GO term enrichment (FDR < 0.05) of positively (purple) and negatively (green) enriched proteins. I. Cluster map of Reactome-annotated pathways. Normalized and standardized intensity values were used as input.

Next, we applied our DVP workflow to microdissect 1000 shapes from each class in triplicate within our defined regions. The resulting protein amount is equivalent to about 100 intestinal enterocytes, and we reached a median of 3,433 unique proteins, with slightly lower numbers for smaller CDX2-stromal cells compared to larger columnar enterocytes (Figure 3D). Correlation analysis of the triplicates revealed high reproducibility of our DVP workflow (Pearson’s R > 0.92). Comparing the three cell types in the three regions in a correlation cluster matrix revealed high proteomic similarity between epithelial cells within and across areas (CDX2++ and CDX2+, Pearson’s R > 0.9). The proteomes of stromal cells (CDX2-) were less similar to the CDX2++ and CDX2+ cells (Pearson’s R < 0.87). However, they were still quite similar across regions (Figure EV2B).

In a principal component analysis (PCA), the three cell types separated along component 1. Stromal, CDX2-cells clustered together independently of the region. In contrast, epithelial, CDX positive cells were separated in component 2, representing high dysplasia regions vs. those with normal epithelium (Figure 3E). To further explore these differences between epithelial and stromal cells at the molecular level, we compared the proteomes of CDX2++ with CDX2-cells of the same region, which resulted in 1,324 significantly differentially expressed proteins (Figure EV2C). Of those, positively upregulated proteins of the stroma were involved in pathways such as ‘extracellular matrix organization’ or ‘collagen binding’ after GO term enrichment analysis, as expected because of the differences in cell types (Figure EV2C).

Based on the correlation analysis and the PCA, there only minor proteomic difference between CDX2++ and CDX2+ cells within and between regions. We therefore continue analysis only on the CDX2++ cells. When comparing CDX2-, stromal cells between the region with high dysplasia and the region with normal glandular architecture, we found no significantly regulated proteins (Figure 3F). In contrast, 70 proteins differed significantly between these regions for CDX2++ cells (Figure 3G). The second most downregulated protein in high dysplasia regions compared to the region with normal epithelium and immune infiltration was ‘deleted in malignant brain tumors 1’ (DMBT1), a known tumor suppressor involved in mucosal immune defense (Park et al., 2018) which our data now ties to the adenoma to carcinoma transition (Figure EV2D).

In a GO term enrichment analysis, the proteins that exhibited the most significant decrease in the high dysplasia region were primarily indicative of a transition towards increased metabolic activity of the glycolytic pathway. In contrast, we observed a decline of proteins involved in the ‘respiratory electron transport’, and a significant increase of proteins like PFKP, PKM and ALDOC that promote anaerobic glycolysis (Figure 3H). When we filtered the proteins for the Reactome-annotated enriched pathways based on identified GO terms and performed hierarchical clustering of normalized and standardized intensities, we found additional members of the aldehyde (ALDH) and alcohol (ADH) dehydrogenase family to be of consistently lower abundance in regions with high dysplasia. The only exception was ALDH1A1 where protein abundance positively correlated with the severeness of dysplasia (Figure 3I). Further, examining ‘complex I biogenesis’ proteins it became evident that nearly all of the detected NDUF family members were less abundant in areas of high and low dysplasia (Figure 3I). These proteins are essential integral components of the NADH-quinone oxidoreductase in the mitochondrial oxidative phosphorylation system, thus DVP directly and in situ captured the metabolic change from oxidative phosphorylation towards anaerobic glycolysis specifically in CDX2++ epithelial cells of high dysplasia regions, also termed “Warburg effect” (Warburg, 1925).

### Protein levels of DMBT1, MARCKS and CD99 stratify CRA cohort

Given our observation that CDX2++ cells most directly reflect the progression from adenoma to cancer, we next applied DVP to all nine representative adenoma specimens to find markers that help stratifying CRAs. (three tissues per C, HDA and NMN groups). We again isolated 1000 individual shapes from areas with a high density of CDX2++ cells and robustly quantified a median of more than 4200 unique proteins per sample. There were 244 ANOVA significant proteins, on which we performed an unsupervised hierarchical clustering (premutation-based FDR <0.01) (Figure 4A, Table EV2). Triplicates as well as outcome groups grouped together, suggesting robust proteomic difference between tumor tissues which were originally combined into the same one ‘high dysplasia’ group. The heat map indicated a main cluster in each of the groups, whose constituent proteins are shown in Figure 4B. Receptor for Activated C Kinase 1 (RACK1, UniProt ID: D6RF23) and cluster of differentiation 99 (CD99) show were among the top five most regulated in cancer and HDA samples (top profile in Figure 4B). Taken together the proteins in this cluster was predominant in pathways for fatty acid and sterol metabolism (Figure 4C). In the middle profile, ribosomal proteins stood out as markedly up in the NMN subset (Figure 4B). Pathway enrichment analysis further highlighted ‘protein targeting to the endoplasmic reticulum’, ‘establishment of protein localization to the endoplasmic reticulum’, and ‘peptide chain elongation’ (Figure 4C). The bottom profile exclusively corresponds to proteins up in the cancer group, with notable colon cancer proteins such as TMEM173 (or STING, Stimulator of Interferon Genes) (An et al., 2019), PARP1 (poly ADP-ribose polymerase-1) (Dziaman et al., 2014; Wang et al., 2017), and RAB25 (Ras-related protein Rab-25) (Agarwal et al., 2009).

**Figure 4.**
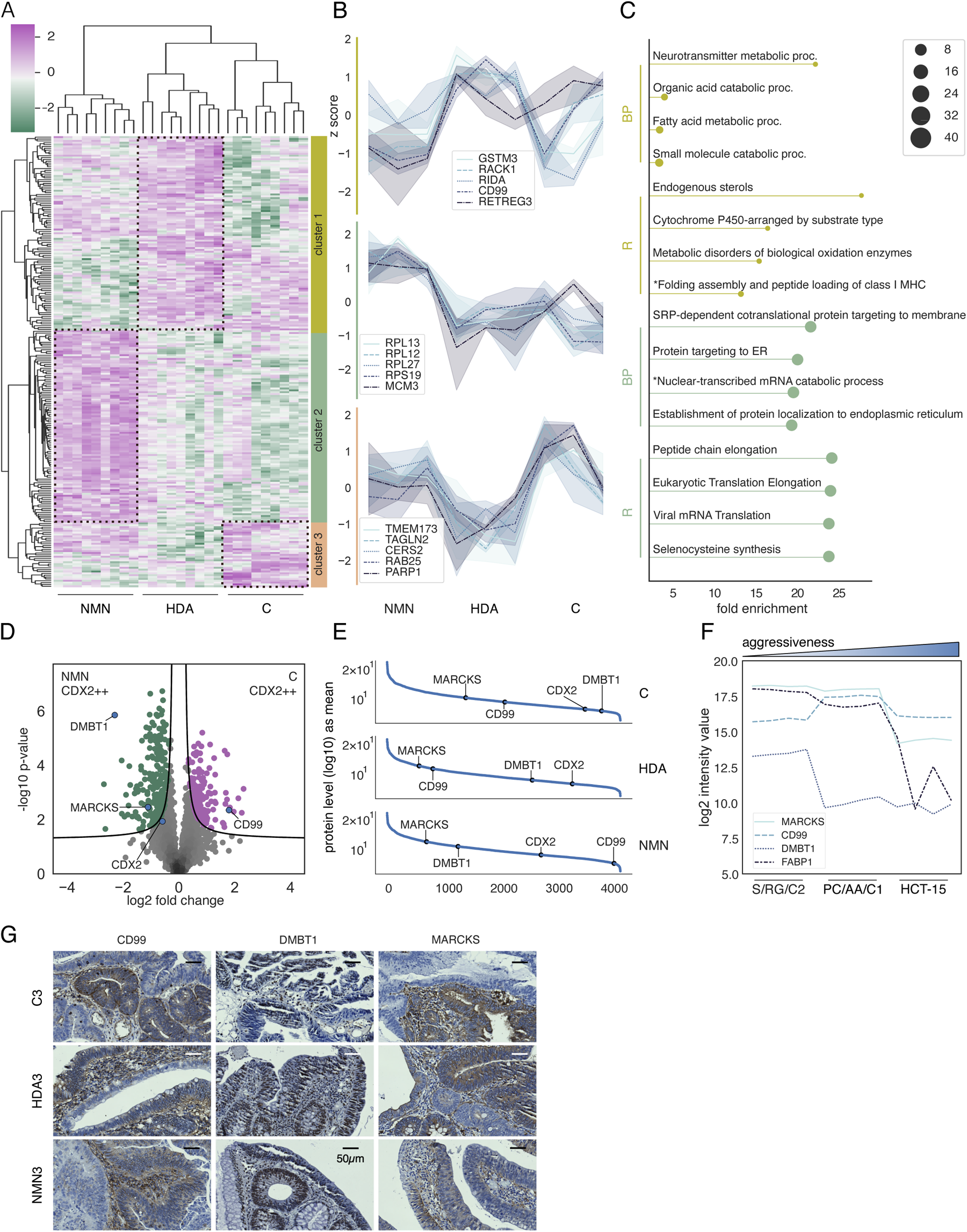
Singly isolated CDX2++ cells from colorectal adenoma tissues from group C, HDA and NMN reveal a potential biomarker set for patient stratification. A. Unsupervised hierarchical clustering of 244 ANOVA significant proteins (premutation-based FDR <0.01, s_0_ = 0.1). B. Line graphs of the top five proteins with the highest ANOVA q value per cluster. C. GO term enrichment analysis of cluster 1 and 2, highlighting Biological Process (BP) and Reactome (R) pathways of proteins with a positive z score. D. Pairwise proteomic comparison of CDX2++ cells comparing the cancer group to NMN (two-sided t-test, FDR <0.01, s_0_ = 0.1). E. Ranked protein abundance of normalized mean intensities of all identified proteins within each CRA group. The potential markers for adenoma classification DMBT1, CD99 and MARCKS are highlighted and labeled. F. Log2 intensity values of these marker proteins across adenoma cell lines S/RG/C2 and PC/AA/C1, and colon carcinoma cell line HCT-15. G. Representative CRA images of IHC staining. Scale bar, 50 µm.

Further, we found myristoylated alanine rich protein kinase C substrate (MARCKS), CD99 and DMBT1 to be of particular interest. These proteins exhibited consistent patterns of enrichment in CDX2++ cells across the nine CRA samples and in particular between the C and NMA group (Figure 4A and D, Figure EV3B and C) similarly to our findings in a single heterogeneous tissue described above (Figure 3G). DMBT1 was the strongest outlier (fold change and significance) that was downregulated in the C group, increasing in abundance in HDA and NMN and in a more indicative manner than our established marker CDX2 (Figure 4E). Conversely, the levels of CD99 were high in groups C and HDA and decreased substantially in NMN (Figure 4E). MARCKS displayed a moderate decrease in abundance from C to NMN, but still statistically significant (Figure 4E). These observations in the rank order plots were mirrored in the assessment of absolute log2-transformed LFQ intensities (Figure EV3E).

Next, we sought to orthogonally investigate DMBT1, CD99 and MARCKS as potential marker set for adenoma stratification. To mimic increasing aggressiveness of tumor cells we evaluated the proteomics profile of these three proteins in two well-established adenoma cell lines S/RG/C2 (Butt et al., 1997) and PC/AA/C1 (Williams et al., 1990, 1991), as well as in the colorectal carcinoma model system HCT-15 (Gonçalves et al., 2022). The levels of our marker set DMBT1, CD99 and MARCKS did correlated with the malignancy status in accordance with our previous results (Figure 4F). To further confirm this, we performed an IHC staining of all adenoma samples. Indeed, DMBT1 and MARCKS exhibited expression patterns consistent with the proteomic findings (Figure 4G). CD99 did not, which may be attributable to its heterogeneous distribution within the stromal and epithelial lining compartments (Figure EV3F).

Taken together, we report MARCKS, CD99, and DMBT1 as potential markers for the transition from adenoma to carcinoma in CDX2++ cells in regions of high dysplasia.

## Discussion

In this paper, we applied the power of Deep Visual Proteomics (DVP) to the important challenge of adenoma stratification beyond current clinical practice. Our goal was to develop and implement a technological framework compatible with routine histological assessments of colorectal adenomas while adding a streamlined, cell-type specific proteomics read out. For our chosen sub-cohort of nine patients representing three outcomes (cancer, high-dysplasia adenoma and non-metachronous neoplasms after ten years). We first generated a deep spectral library by DDA measurements of extensively fractionated bulk samples of each of these samples, leading to an in-depth CRA proteome, a valuable resource of more than 12,000 unique proteins. We found minimal significant differences between the outcome groups due to insufficient quantitative accuracy in these fractionated, DDA data. This was partially alleviated by bulk DIA measurements that used the deep spectral libraries acquired by DDA. However, to move beyond global features such as enrichment of proteins involved in extracellular matrix (ECM) organization or cell adhesion in C, suggesting structural changes which are in line with enhanced tumor cell migration and invasion (Poltavets et al., 2018; Winkler et al., 2020), we needed to move to a cell-type specific approach.

We then turned to Deep Visual Proteomics to investigate intra-tumor heterogeneity. Starting with one particularly heterogeneous adenoma sample, we defined regions with increasing levels of dysplasia. We further defined three cell types based on the staining of CDX2, a common marker for high grade dysplasia, namely CDX2++, CDX2+ and CDX2-. This immediately revealed differences in energy metabolism across different regions, which were attributable to CDX2++ epithelial cells. These notably increased proteins associated with glycolysis and decreased mitochondrial complex I proteins in regions with high dysplasia (Vander Heiden et al., 2009), a direct, in situ and cell-type specific observation of the Warburg effect. DVP assigned this change specifically to highly dysplastic areas within the tumor, rather than being observed throughout the entire adenoma polyp. In addition, we observed a reduced expression of detoxification enzymes in high dysplasia areas which may cause intensified carcinogenesis (Lindahl, 1992). Specifically, ALDH1A1 levels increased in CDX2++ cells in regions of high dysplasia compared to other members of the aldehyde dehydrogenase protein family. This protein primarily participates in the oxidation process of retinaldehyde to retinoic acid, which promotes cell proliferation and inhibits apoptosis through the action of the transcription factor and proto-oncogene c-MYC (Tomita et al., 2016; Zanoni et al., 2022). Differential expression of ALDH and AHD family members has been controversially discussed as either promoting or inhibiting cancer progression. Our data support that ALDH1A1, which has already been proposed as a prognostic maker for early invasiveness of cancer (Althobiti et al., 2020; Yang et al., 2014), is clearly upregulated in regions of high dysplasia.

Given these results, we exclusively focused on proteomic investigation of CDX2++ epithelial cells across all nine CRA individuals. The CDX2++ proteomes clustered not only by replicates of the same patient, but importantly by outcome, suggesting the existence of protein signatures associated with the adenoma to carcinoma transition. Specifically, we found the proteins DMBT1, MARCKS and CD99 to be associated with each recurrence outcome. Interestingly DMBT1, which has been reported to act as tumor suppressor in various cancer types (Mori et al., 1999; Somerville et al., 1998), exhibited the strongest negative fold change. DMBT1 expression has primarily been observed in cells of the immune system and epithelial linings (Mollenhauer et al., 2000; Mori et al., 1999), driving differentiation but also carcinogenesis when absent, which makes it promising candidate for driving adenoma-carcinoma progression. MARCKS was another interesting protein candidate as it was solely significantly enriched in the cancer group. It is a major target of PKC and has repeatedly been implicated in tumor progression (Fong et al., 2017). As a plasma membrane-tethered protein that shuttles into the cytosol upon phosphorylation, MARCKS functions as a regulator of cellular signaling, affecting sensitivity to programmed cell death and cell adhesion through various pathways(Fong et al., 2017). Although the role of MARCKS in the development of cancers remains a topic of debate, it may suppress cell growth in colorectal cancer (Bickeböller et al., 2015; Fong et al., 2017; Rombouts et al., 2013). Like DMBT1, spatial proteomics implicates MARCKS as being involved in the transition towards cancer. Lastly, CD99 is discussed as an onco-suppressor or an oncogene (Manara et al., 2018). It is involved in a diverse range of molecular processes, including the reduction of miR34a/Notch/NF-κB and MAPK pathways (Byun et al., 2006; Seol et al., 2012; Ventura et al., 2016). In our study, CD99 exhibited significant upregulation in high grade-dysplasia and cancer outcome groups while it was undetectable in the NMN group, suggesting it as a driver for cell migration and tumor invasiveness (Byun et al., 2006; Seol et al., 2012).

In conclusion, our findings offer insights into the intra- and inter-individual biology of adenomas that later manifest different outcomes. Cell-type specific spatial proteomics highlighted the most informative cell types and regions and revealed the existence of protein patterns implicated in the transition through the adenoma to carcinoma sequence. It also identified a subset of potential markers or molecular drivers that could be investigated further with the purpose of functionally characterizing cancer progression or developing a panel for risk stratification.

## Materials and Methods

### Study design and sample collection

This is a retrospective study, based on adenomas resected between 2006-2011 at Odense University Hospital and formalin-fixed and paraffin-embedded for preservation. Samples were provided by the Danish pathology data bank (Patobank) and anonymized, after reevaluation of high-grade dysplasia status of each poly by a pathologist. Using clinical data on disease recurrence, we categorized nine CRAs into three groups of three: group C (cancer), HDA (high-grade dysplasia adenoma), and NMN (non-metachronous neoplasms).

### Ethical permission

The experimental design including the Deep Visual Proteomics workflow was approved by the National Committee on Health Research Ethics (j.nr. 2112779), and a waiver for obtaining informed consent was granted (as per section 10, subsection 1 of the Committee Act).

### Immunohistochemistry

A detailed protocol for FFPE tissue mounting and staining on membrane PEN slides 1.0 (Zeiss, 415190-9041-000) is provided in our original DVP article^1^.

The antibodies we have used to either characterize HG dysplasia in CRAs, or for marker validation were always counterstained with Mayer’s hematoxylin and optimized to the following working concentrations (Table 1).

**Table 1.**
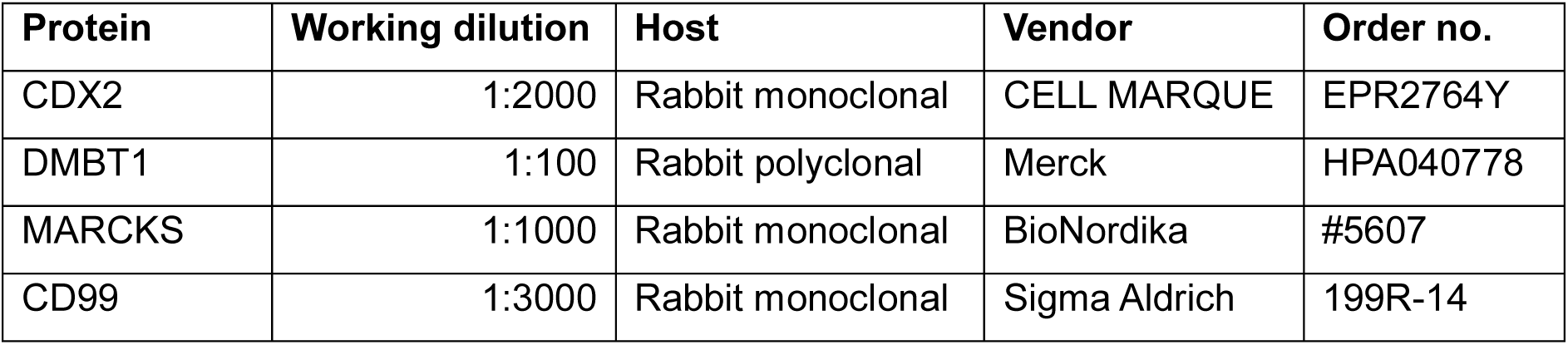
List of antibodies.

### Immunofluorescence staining

The tissue sections underwent deparaffinization and hydration through three cycles of xylene and decreasing ethanol concentrations from 99.6% to 70%. Antigen retrieval was achieved by immersing sections in 10 mM citrate buffer (pH 6.0) at 90°C for 20 minutes. Subsequently, tissues were blocked with 5% BSA for 1 hour at room temperature. Following overnight incubation at 4°C with anti-CDX2 (235R-15, Cell Marque; 1:1000) antibodies, slides were washed and then incubated with Alexa Fluor® 568 goat anti-rabbit antibody (A-11036, Invitrogen; 1:1000) for one hour at room temperature. After rinsing, slides were further incubated with anti-CD3 antibody (A0452, Agilent; conjugated with DyLight® 488 (ab201799, Abcam); 1:300) and anti-CD20 (50-0202-80, Invitrogen; 1:100) overnight at 4°C. DAPI was used for counterstaining, and slides were mounted with Thermo Fisher Diamond Antifade mounting media before examination under an AxioScan7 microscope.

### Laser microdissection

After reference point alignment at the LMD7 (Leica) microscope, shape contours were imported for semi-automated laser microdissection at the following setting: power 32, aperture 1, speed 20, final pulse −1, head current 42%, pulse frequency 2,600 and offset 180/220. All experiments were controlled with the LMD software v8. 1000 shapes were cut and sorted into 384-well plates (Eppendorf 0030129547), avoiding the collection in the outermost rows and columns. After microdissection, plates were sealed, centrifugated at 1,000g for 10 min and then frozen at −20 °C until further processing.

### High-resolution microscopy

IHC-stained FFPE tissue sections of 2.5 µm thickness were scanned with the Zeiss Axio ScanZ.1 microscope. With a x20, 0.8 NA dry objective, widefield images were acquired using a VIS LED light source and captured by a CCD Hitachi HV-F202CLS camera. Dependent on given tissue irregularities on PEN membrane slides, the z-stack configurations were set to five to fifteen slices and a regular interval of 1.50 µm to guarantee sample coverage and optimal focus. Having ‘EDF active’ (Extended depth of focus) checked during acquisition, a 2D-projection based on maximum intensity values was created and used to generate a stitched tissue image (Zeiss ZEN 2.6, blue edition) for further image processing.

### Cell segmentation and classification

The DVP approach in this investigation focuses on standardized IHC and H&E staining, where cellular borders are often indistinct. We therefore utilized a deep neural network in BIAS for cell segmentation that was trained on a ‘generic nuclei data set’, and set a fixed cutting-offset of 2 µm for including the cytoplasm in proteome analysis. To classify epithelial CDX2 cells, we exported the image feature matrix from BIAS to a custom Jupyter Notebook. After removing outliers via a 5% z-scored intensity cutoff, cells were classified into four groups using nuclei RGB intensities and morphologies. The processed matrix was returned to BIAS for contour export and laser microdissection.

### MS sample preparation

One thousand dilated nuclei contours, equivalent to about 100 colonic epithelial cells (BNID 111216), were automatically excised with the cutting offset and pooled into a 384-well plate (Eppendorf, 0030129547). For each defined adenoma tissue area, we collected triplicates. All MS sample preparation steps and buffers were replicated from the original DVP paper (Mund et al., 2022), but semi-automated by using the Agilent Bravo liquid handling robot. After digestion, samples were vacuum dried and stored at −20°C, or resuspended in 20 µL Evosep buffer A (0.1% formic acid v/v) for direct Evotip pure loading (www.evosep.com).

### High-pH reverse-phase fractionation

We used high-pH reverse-phase fractionation to create a deep spectral library of CRA material for subsequent used in Data Independent Acquisition (DIA). To this end, we utilized our automated Opentrons platform for fraction collection coupled to a nanoflow HPLC (EASY-nLC 1000 system, Thermo Fisher Scientific). For this bulk analysis, FFPE adenoma tissue was scraped from the PEN glass slide and enzymatically digested proteins for MS analysis. Peptides were then separated on an analytical column (250 µm x 30 cm, 1.9 µm, PepSep™, Bruker Daltonics) by a 100 min gradient with an exit-valve switch every 30 seconds and concatenated into 48 fractions.

### LC-MS

To ensure minimal loss of peptides, we directly queued our samples after Evotip loading for LC-MS analysis. Acquisition was performed on a timsTOF instrument (Bruker Daltonics, timsTOF SCP) coupled with a Evosep One system. Using the 30 Sample Per Day (SPD) method (www.evosep.com), samples were separated on an analytical column (150 µm x 15 cm, 1.5 µm; PepSep™, Bruker Daltonics), and a 10 µm emitter operated inside a captive nano-electrospray ion source (Bruker Daltonics). We set the ion accumulation and elution time in the TIMS tunnel to 100 ms. Samples were either measured in data-dependent (ddaPASEF) or data-independent (diaPASEF) modes, with parameters like m/z-range, ion mobility, and PASEF cycles following the specifications in Brunner et al.(Brunner et al., 2022). Quality control samples and our deep CRA library were measured in ddaPASEF. DiaPASEF was used for low input FFPE DVP samples.

### MS data analysis

Bruker timsTOF ddaPASEF raw files were analyzed with AlphaPept (version 0.4.1) (Strauss et al., 2021) using standard settings (https://mannlabs.github.io/alphapept/settings.html). For our 432 deep CRA samples, each of the 48 fractions was assigned to a patient in AlphaPept. Using the same DDA files, we created a project-specific CRA library in MSFragger (v18.0) with 178,274 precursors and 12,389 unique protein groups, excluding cysteine carbamidomethylation as fixed modification (Kong et al., 2017b). Its IonQuant (v1.8.9) and Philosopher (v4.2.2) modules handled quantification and False Discovery Rate (FDR) correction, respectively. Low input DVP samples in diaPASEF mode were analyzed in DIA-NN (v1.8.1) (Demichev et al., 2020, 2022), using a library-based approach against the UniProt database with isoforms (2019, UP000005640_9606). Settings included trypsin specificity with one missed cleavage, 1% precursor FDR, 15 ppm accuracy, and enabled ‘match between runs’. N-terminal methionine excision, methionine oxidation and N-terminal acetylation were left checked, and maximal 2 variable modification were allowed.

### Spectral library generation

For low input DVP sample analysis which were acquired in diaPASEF, we utilized our project-specific deep colorectal adenoma library created using FragPipe^2^ (version 17.1, incorporating MSFragger 3.4^3,4^, Philosopher 4.1.1^5^, Python 3.9.7, and EasyPQP 0.1.25, available at https://github.com/grosenberger/easypqp). While default parameters were largely maintained, adjustments were made to set the precursor mass tolerance between −20 and 20 ppm and the fragment mass tolerance at 20 ppm. The resulting data tables were subjected to a 1% false discovery rate (FDR) filter using Percolator and ProteinProphet options in FragPipe.

### Bioinformatic analysis

Fractionation data acquired in ddaPASEF was analyzed with AlphaPept to obtain a deep proteome coverage of each tumor bulk sample. The resulting output table was then imported into Perseus (Tyanova et al., 2016), and filtered for protein groups with 70% of quantitative values present ‘in at least one group’ (C, HDA, NMN). DIA-NN output tables were similarly processed in Perseus. Before statistical testing, missing values were imputed based on a normal distribution (width = 0.3; downshift = 1.5). To correct for multiple hypothesis testing in pairwise proteomic comparisons (two-sided unpaired t-test), we applied a permutation-based false discovery rate (FDR) of either 5% or 1%, as specified in the figure legends. We corrected a multi-sample ANOVA for a 1% false discovery rate (FDR). Gene Set Enrichment Analysis (GSEA) was done in Python 3.9.7 (https://github.com/zqfang/GSEApy, v1.0.4). For visualization, we used the Python libraries NumPy (v1.20.3), Pandas (v1.3.4), Matplotlib (v3.4.3), and Seaborn (v0.12.2). Gene ontology (GO) term enrichment analysis was performed online utilizing the ShinyGo application (http://bioinformatics.sdstate.edu/go/), v0.77).

## Acknowledgements

The authors would like to thank L. Drici (NNF CPR Proteomics Program) and J. Madsen (NNF CPR Mass Spectrometry Platform, University of Copenhagen) for technical assistance. We acknowledge P. Hernandez-Varas and Richard Denis Maxime De Mets form the Core Facility of Integrated Microscopy for microscopy (CFIM) support, and C. Greb and F. Schlaudraff from Leica for technical support. We thank F.Mundt, L. Schweizer, J. Wang, M. Thielert, F. Coscia and P. Skowronek for fruitful discussions.

## CRediT Author contributions

S. Kabatnik, (Conceptualization: Lead; Investigation: Lead; Writing – original draft: Lead).

F. Post, (Conceptualization: Supporting; Data curation: Supporting).

L. Drici, PhD, MS (Investigation: Supporting).

A.S. Bartels, (Investigation: Supporting).

M.T. Strauss, PhD (Conceptualization: Supporting; Writing – editing: Supporting).

X. Zheng, PhD (Investigation: Supporting).

G. I. Madsen, MD/PhD (Investigation: Supporting).

A. Mund, PhD (Writing – review: Supporting).

F.A. Rosenberger, PhD (Writing – review & editing: Lead).

J.M.A. Moreira, PhD (Conceptualization: Equal; Supervision: Equal; Resources: Supporting; Project administration: Equal; Writing – review and editing: Equal).

M. Mann, PhD (Conceptualization: Equal; Supervision: Equal; Resources: Lead; Project administration: Equal; Funding acquisition: Lead; Writing – review and editing: Equal).

## Disclosure and competing interest statement

M. Mann is an indirect investor in Evosep Biosystems. The other authors declare have no conflicts of interest with the contents of this article.

## The Paper Explained

### Problem

Colorectal cancer (CRC) is the third most prevalent cancer worldwide and is a leading cause of cancer-related mortality. Screening for CRC has been shown to effectively reduce mortality, as adenomas can be detected, removed, and used for patient risk classification. Although various factors are considered for patient stratification, the detection of high-grade dysplasia in a polyp often leads to the advice of annual follow-up colonoscopies. Since only less than 10% of these patients actually progress to cancer, it would be useful to improve risk stratification.

### Results

We have recently developed a spatial workflow that combines digital pathology, laser microdissection of single cell types and ultra-sensitive proteomics (DVP). DVP seamlessly integrated with current pathology workflows and equipment, identifying deleted in malignant brain tumors 1 (DMBT1), myristoylated alanine rich protein kinase C (MARCKS), and cluster of differentiation 99 (CD99) correlated with disease recurrence history, making them potential markers of risk stratification. The spatial and cell-type specific capabilities of DVP uncovered a metabolic switch towards anaerobic glycolysis in areas of high dysplasia, which was specific for cells with high CDX2 expression.

### Impact

Our research provides an in depth description of the proteomic alterations in colorectal adenomas. It highlights pathways for future studies on the progression and invasion mechanisms of colorectal cancer, potentially leading to novel therapeutic strategies.

## Funding

This work is supported financially by the Novo Nordisk Foundation (grant NNF14CC0001), the Max Planck Society and the Sawmill Owner Jeppe Juhl and Wife Ovita Juhl Memorial Foundation. Additionally, S. Kabatnik and F. Post were supported by the Novo Nordisk Foundation grant NNF20SA0035590 and NNF0069780.

## Data availability

The proteomics raw data have been submitted to the ProteomeXchange Consortium through the PRIDE partner repository (https://www.ebi.ac.uk/pride/) with the identifier PXD046999. Image raw data will be provided upon request by contacting Sonja Kabatnik at.

## Notes

### Competing Interest Statement

Matthias Mann is an indirect investor in Evosep Biosystems. The other authors declare have no conflicts of interest with the contents of this article.

