## Supplementary Figures for "Spatial characterization and stratification of colorectal adenomas by Deep Visual Proteomics"

| Group | Age range | Sex | Adenoma localization | Recurrence interval | Relapse type | Tumor type |
|---|---|---|---|---|---|---|
| NMA1 | 56-60 | Male | Rectum | - | - | - |
| NMA2 | 66-70 | Male | Rectum | - | - | - |
| NMA3 | 76-80 | Male | Rectum | - | - | - |
| HDA1 | 71-75 | Male | Rectum | 1 year | HG adenoma | Tubulovillous |
| HDA2 | 71-75 | Male | Rectum | 1 year | HG adenoma | Tubulovillous |
| HDA3 | 81-85 | Male | Rectum | 1 year | HG adenoma | Tubulovillous |
| C1 | 61-65 | Male | Rectum | 1 year | Adenocarcinoma | Tubulovillous |
| C2 | 81-85 | Male | Rectum | 1 year | Adenocarcinoma | Tubulovillous |
| C3 | 76-80 | Male | Colon | 1 year | Adenocarcinoma | Tubulovillous |

**Table EV1 Colorectal adenoma subcohort overview.**

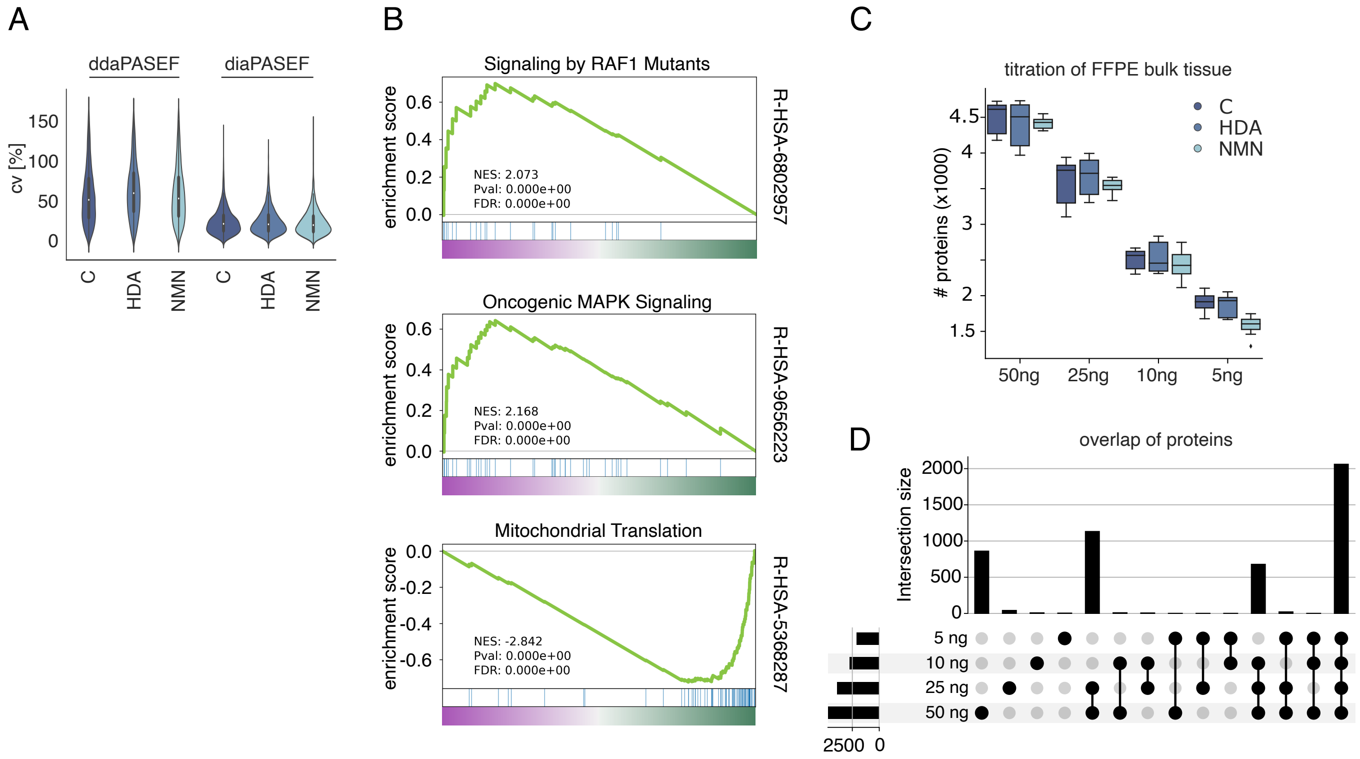
A
ddaPASEF
diaPASEF
cv [%]
150
100
50
0
C
HDA
NMN
C
HDA
NMN
B
Signaling by RAF1 Mutants
enrichment score
NES: 2.073
Pval: 0.000e+00
FDR: 0.000e+00
R-HSA-6802957
Oncogenic MAPK Signaling
NES: 2.168
Pval: 0.000e+00
FDR: 0.000e+00
R-HSA-9656223
Mitochondrial Translation
NES: -2.842
Pval: 0.000e+00
FDR: 0.000e+00
R-HSA-5368287
C
titration of FFPE bulk tissue
### proteins (x1000)
C
HDA
NMN
50ng
25ng
10ng
5ng
D
overlap of proteins
Intersection size
5 ng
10 ng
25 ng
50 ng
2500
0

**Figure EV1 Biological and comparative analysis of DIA and DDA acquired data from bulk CRA FFPE samples.**

A. Gene Set Enrichment Analysis (GSEA) of ddaPASEF acquired data, originating from fractionated bulk FFPE tissue samples.

B. Number of unique proteins at decreasing FFPE bulk lysate injections (50ng, 25 ng, 10 ng and 5 ng), measured in DIA and quantified by library-based DIA-NN.

C. Overlap of proteins across different injection amounts.

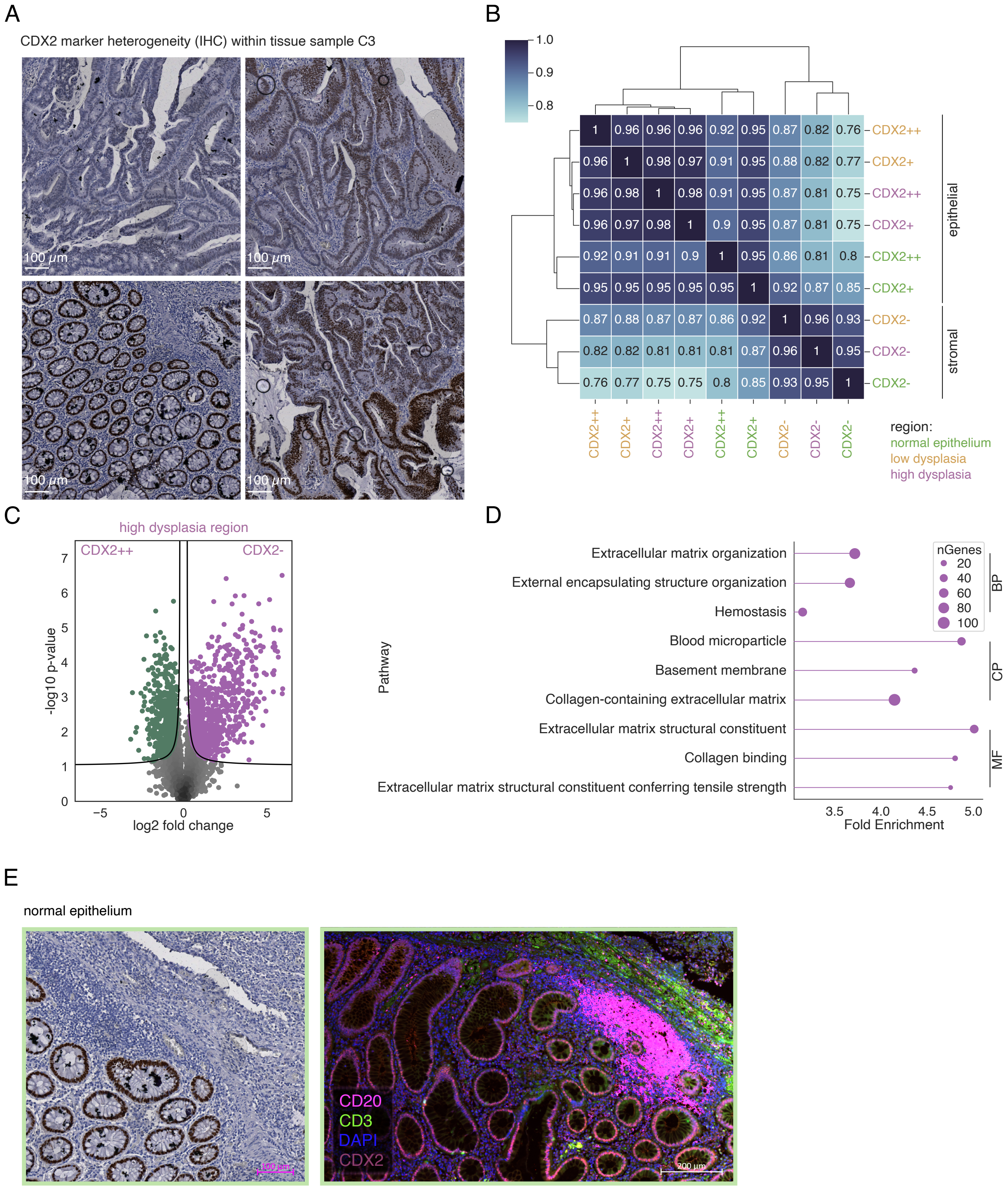
A
CDX2 marker heterogeneity (IHC) within tissue sample C3
100 μm
B
1.0
0.9
0.8
1 0.96 0.96 0.96 0.92 0.95 0.87 0.82 0.76 CDX2++
0.96 1 0.98 0.97 0.91 0.95 0.88 0.82 0.77 CDX2+
0.96 0.98 1 0.98 0.91 0.95 0.87 0.81 0.75 CDX2++
0.96 0.97 0.98 1 0.9 0.95 0.87 0.81 0.75 CDX2+
0.92 0.91 0.91 0.9 1 0.95 0.86 0.81 0.8 CDX2++
0.95 0.95 0.95 0.95 0.95 1 0.92 0.87 0.85 CDX2+
0.87 0.88 0.87 0.87 0.86 0.92 1 0.96 0.93 CDX2-
0.82 0.82 0.81 0.81 0.81 0.87 0.96 1 0.95 CDX2-
0.76 0.77 0.75 0.75 0.8 0.85 0.93 0.95 1 CDX2-
epithelial
stromal
CDX2++ CDX2+ CDX2++ CDX2+ CDX2++ CDX2+ CDX2- CDX2- CDX2-
region:
normal epithelium
low dysplasia
high dysplasia
C
high dysplasia region
CDX2++
CDX2-
-log10 p-value
log2 fold change
D
Extracellular matrix organization
External encapsulating structure organization
Hemostasis
Blood microparticle
Basement membrane
Collagen-containing extracellular matrix
Extracellular matrix structural constituent
Collagen binding
Extracellular matrix structural constituent conferring tensile strength
Pathway
nGenes
20
40
60
80
100
BP
CP
MF
Fold Enrichment
E
normal epithelium
CD20
CD3
DAPI
CDX2
200 μm

**Figure EV2 Colorectal adenoma heterogeneity.**

A. IHC staining against the CDX2 marker protein of one particularly heterogenous colorectal adenoma (CRA) tissue (C3). Scale bar, 100 µm.

B. Correlation matrix of log2 normalized and imputed intensity values of diaPASEF-acquired low-input DVP samples.

C. Pairwise proteomic comparison between cell classes CDX2++ (epithelial) and CDX2- (stromal), both laser microdissected from our pre-defined highly dysplastic adenoma region. Significantly enriched proteins are colored and displayed above the black lines (two-sided t-test, permutation-based FDR <0.05, $s_0$ = 0.1).

D. GO term enrichment (FDR <0.05) of significantly positive protein hits, showing pathways included in 'Biological Process' (BP), 'Cellular Process' (CP), and 'Molecular Function' (MF).

E. Area showing normal glandular architecture of CRA C3. Left: IHC against CDX2 and hematoxylin counterstaining. Right: Immunofluorescence staining against B lymphocytes (CD20), T lymphocytes (CD4) a nuclear staining (DAPI), and CDX2.

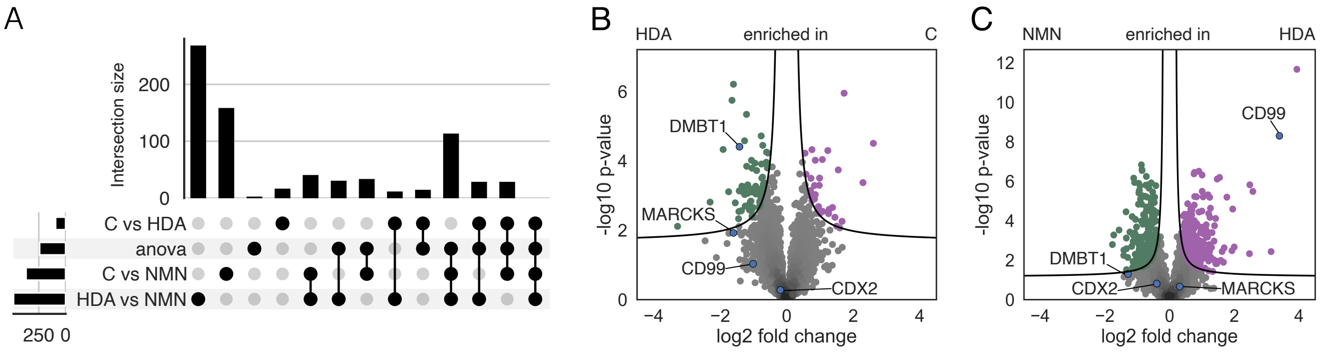


D

**C vs NMN**

| Enriched in C | | Enriched in NMN | | -log10 p-value | |
|---|---|---|---|---|---|
| Protein Group | Gene Name | Protein Group | Gene Name | Protein Group | Gene Name |
| P07148 | FABP1 | O95620 | DUS4L | P26373 | RPL13 |
| Q9ULW5 | RAB26 | Q9BYZ8 | REG4 | Q92979 | EMG1 |
| Q9UGM3-3 | DMBT1 | Q8NFC6 | BOD1L1 | P62263 | RPS14 |
| Q9NQU5-2 | PAK6 | A0A087WWM1 | MUC1 | P05388 | RPLP0 |
| P80188 | LCN2 | Q01995 | TAGLN | P30050 | RPL12 |
| Q96JA1-2 | LRIG1 | P14209 | CD99 | A8MUS3 | RPL23A |
| A6NHN0 | OTOL1 | Q5T7N2 | L1TD1 | Q8WXI4-2 | ACOT11 |
| A8K7I4 | CLCA1 | P18065 | IGFBP2 | P39019 | RPS19 |
| Q53HL2 | CDCA8 | Q6P179 | ERAP2 | Q9UGM3-3 | DMBT1 |
| Q8IYS1 | PM20D2 | P17661 | DES | A8K7Q2 | HSPA8 |
| Q9Y2I7 | PIKFYVE | O75891-3 | ALDH1L1 | Q02878 | RPL6 |
| A0A0D9SEJ5 | FAM120B | D6RF23 | RACK1 | Q92820 | GGH |
| P12277 | CKB | Q92526-3 | CCT6B | P62269 | RPS18 |
| P16444 | DPEP1 | U3KQE2 | CAPNS1 | P12236 | SLC25A6 |
| P01859 | IGHG2 | P61764 | STXBP1 | P61247 | RPS3A |
| Q9H300 | PARL | P30260-2 | CDC27 | P40429 | RPL13A |
| A0A075B7D8 | IGHV3OR15-7 | P48163 | ME1 | P62424 | RPL7A |
| Q6P4A8 | PLBD1 | P84074 | HPCA | Q3YEC7 | RABL6 |
| Q9NRD8 | DUOX2 | C9JZL8 | MYADM | Q9BXR0 | QTRT1 |
| Q9UPV0 | CEP164 | Q9HC84 | MUC5B | Q14240-2 | EIF4A2 |
| Q5BJH7-2 | YIF1B | Q9Y4H2 | IRS2 | P62899 | RPL31 |
| P26447 | S100A4 | P49023-4 | PXN | Q86VR2 | RETREG3 |
| Q07954 | LRP1 | Q8N0U8 | VKORC1L1 | P61313 | RPL15 |
| Q96B21 | TMEM45B | Q69YN4 | VIRMA | D6R9L0 | RACK1 |
| P51553 | IDH3G | Q9BZF9 | UACA | P62753 | RPS6 |
| O15120 | AGPAT2 | Q86TX2 | ACOT1 | P32969 | RPL9 |
| P02786 | TFRC | Q7L5N7 | LPCAT2 | P36578 | RPL4 |
| Q9P219 | CCDC88C | P08758 | ANXA5 | P18124 | RPL7 |
| Q68CZ6 | HAUS3 | P02760 | AMBP | P46776 | RPL27A |
| P49407 | ARRB1 | Q96SQ9 | CYP2S1 | O60506 | SYNCRIP |

**C vs HDA**

| Enriched in C | | Enriched in HDA | | -log10 p-value | |
|---|---|---|---|---|---|
| Protein Group | Gene Name | Protein Group | Gene Name | Protein Group | Gene Name |
| Q8WWA0 | ITLN1 | Q7Z333-4 | SETX | Q7LG56 | RRM2B |
| A0A087WWM1 | MUC1 | D6RF23 | RACK1 | P18065 | IGFBP2 |
| P18065 | IGFBP2 | O43704 | SULT1B1 | P21266 | GSTM3 |
| Q01995 | TAGLN | P18827 | SDC1 | P52758 | RIDA |
| C9JZL8 | MYADM | P21266 | GSTM3 | O00273 | DFFA |
| A0A0B4J259 | LYZ | Q7LG56 | RRM2B | P10253 | GAA |
| P06731 | CEACAM5 | Q9NQU5-2 | PAK6 | Q8WWA0 | ITLN1 |
| P09382 | LGALS1 | P09467 | FBP1 | P09496-2 | CLTA |
| Q969L2 | MAL2 | P42765 | ACAA2 | P29966 | MARCKS |
| Q8NFJ5 | GPRC5A | P22570-7 | FDXR | O43704 | SULT1B1 |
| Q86WV6 | TMEM173 | P29966 | MARCKS | P57735 | RAB25 |
| Q8WUY3-2 | PRUNE2 | P45954 | ACADSB | Q86WV6 | TMEM173 |
| P08758 | ANXA5 | Q02318 | CYP27A1 | P21912 | SDHB |
| Q6EEV4 | POLR2M | Q07954 | LRP1 | Q7Z2K6 | ERMP1 |
| O94886 | TMEM63A | P19801-2 | AOC1 | P09874 | PARP1 |
| P21333-2 | FLNA | P10253 | GAA | P08758 | ANXA5 |
| Q7Z4F1 | LRP10 | P35244 | RPA3 | Q5T6V5 | C9orf64 |
| B8ZZT4 | VAMP8 | O60218 | AKR1B10 | O75947 | ATP5H |
| P18031 | PTPN1 | Q5GFL6-2 | VWA2 | O14936 | CASK |
| Q5T6V5 | C9orf64 | P52758 | RIDA | O94819 | KBTBD11 |
| Q9Y223-2 | GNE | P02747 | C1QC | P29218-3 | IMPA1 |
| P00966 | ASS1 | Q9UHK6-5 | AMACR | Q16762 | TST |
| Q15738 | NSDHL | Q9BSE5 | AGMAT | P02747 | C1QC |
| P57735 | RAB25 | Q96EL3 | MRPL53 | P61457 | PCBD1 |
| P05783 | KRT18 | P49748 | ACADVL | Q9UIJ7 | AK3 |
| Q96G23 | CERS2 | Q07021 | C1QBP | C9JZL8 | MYADM |
| F8WD26 | LMO7 | P30711 | GSTT1 | P25815 | S100P |
| Q9Y4H2 | IRS2 | Q96CM8-2 | ACSF2 | P37802 | TAGLN2 |
| P37802 | TAGLN2 | Q96GK7 | FAHD2A | P17152 | TMEM11 |
| P09874 | PARP1 | Q9H061 | TMEM126A | P21333-2 | FLNA |

**HDA vs NMA**

| Enriched in HDA | | Enriched in NMN | | -log10 p-value | |
|---|---|---|---|---|---|
| Protein Group | Gene Name | Protein Group | Gene Name | ProteinGroup | Gene Name |
| D6RF23 | RACK1 | P16444 | DPEP1 | P14209 | CD99 |
| P14209 | CD99 | Q68CZ6 | HAUS3 | P46776 | RPL27A |
| Q5MY95 | ENTPD8 | Q9NRD8 | DUOX2 | P61353 | RPL27 |
| U3KQE2 | CAPNS1 | P26447 | S100A4 | P18124 | RPL7 |
| P10109 | FDX1 | P07148 | FABP1 | Q92905 | COPS5 |
| Q9Y4C4 | MFHAS1 | Q16666-2 | IFI16 | Q6YN16 | HSDL2 |
| P01877 | IGHA2 | Q14657 | LAGE3 | Q9HBL7 | PLGRKT |
| Q8IVG5 | SAMD9L | P02786 | TFRC | H3BPR2 | NME3 |
| P02461 | COL3A1 | A0A0D9SEJ5 | FAM120B | P04075 | ALDOA |
| Q9Y3Y2-3 | CHTOP | P80188 | LCN2 | G3V2F5 | PRMT5 |
| Q9H008 | LHPP | Q9HCE7-2 | SMURF1 | P40429 | RPL13A |
| Q92526-3 | CCT6B | Q9UPV0 | CEP164 | P21266 | GSTM3 |
| P48163 | ME1 | Q9UHX1-6 | PUF60 | P25205 | MCM3 |
| P21695-2 | GPD1 | Q96EL2 | MRPS24 | P26373 | RPL13 |
| G3V2F5 | PRMT5 | P05534 | HLA-A | P61313 | RPL15 |
| P21266 | GSTM3 | Q6DN03 | HIST2H2BC | P15531-2 | NME1 |
| Q96T58 | SPEN | Q9Y2I7 | PIKFYVE | P10109 | FDX1 |
| O75891-3 | ALDH1L1 | Q92820 | GGH | P49207 | RPL34 |
| P01591 | JCHAIN | P40429 | RPL13A | Q13310-2 | PABPC4 |
| Q96CD0 | FBXL8 | Q96RQ1 | ERGIC2 | P50914 | RPL14 |
| P62306 | SNRPF | P10412 | HIST1H1E | P39019 | RPS19 |
| P21397 | MAOA | P25205 | MCM3 | C9JNW5 | RPL24 |
| H0YD16 | TOR1AIP1 | F5H124 | MAGOHB | P62851 | RPS25 |
| Q5T7N2 | L1TD1 | Q9BTT0-3 | ANP32E | P51572-2 | BCAP31 |
| P07902-2 | GALT | P07195 | LDHB | U3KQE2 | CAPNS1 |
| Q9BRQ6 | CHCHD6 | Q8NEW0 | SLC30A7 | P37802 | TAGLN2 |
| Q9BRF8 | CPPED1 | Q86WV6 | TMEM173 | P62424 | RPL7A |
| Q9H299 | SH3BGRL3 | Q99525 | HIST1H4G | Q96CM8-2 | ACSF2 |
| Q9BWH2 | FUNDC2 | Q9UIG0-2 | BAZ1B | Q14566 | MCM6 |
| P11217 | PYGM | Q9BWD1 | ACAT2 | P33993 | MCM7 |

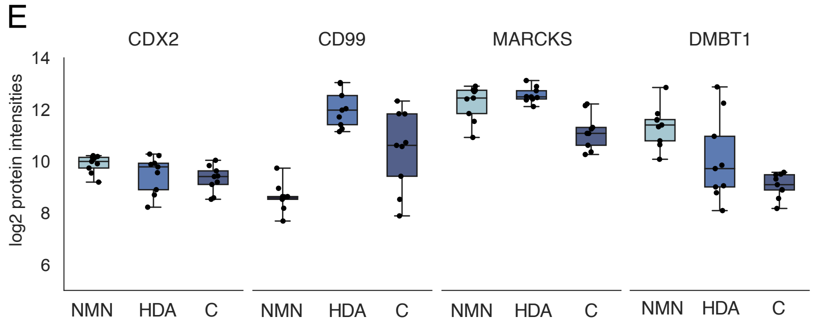


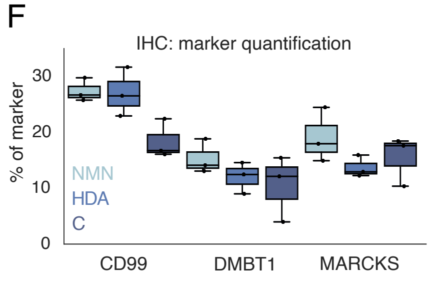

**Figure EV3 Proteomic overview from singly isolated CDX2++ cells from our colorectal adenoma subcohort.**

A. Overlap of significantly enriched proteins after pairwise proteomic comparison of CDX2++ cells (two-sided t-test, permutation-based FDR <0.01, $s_0$ = 0.1) across CRA tissues.

B, C. Pairwise proteomic comparison between group HDA and C, and NMN and HDA (two-sided t-test, permutation-based FDR <0.01, $s_0$ = 0.1).

D. List of significantly enriched proteins between groups C, HDA and NMN after pairwise proteomic comparison (two-sided t-test, permutation-based FDR <0.01, $s_0$ = 0.1). Top 30 significantly enriched proteins are presented in a descending sequence, ordered according to their -log10 p-value.

E. Log2-transformed LFQ intensities of marker proteins CD99, DMBT1 and MARCKS.

F. IHC signal quantification of marker proteins CD99, DMBT1 and MARCKS.
